# SydneyMTL: Interpretable Multi-Task Learning for Complete Sydney System Assessment in Gastric Biopsies

**DOI:** 10.64898/2026.02.17.26346304

**Authors:** Won Chan Jeong, Ho Heon Kim, Yuri Hwang, Gisu Hwang, Kyungeun Kim, Young Sin Ko

## Abstract

The Updated Sydney System (USS) provides a standardized framework for grading gastritis and stratifying gastric cancer risk. However, subjective observer variability and labor-intensive workflows impede its routine clinical use. To address these challenges, we developed SydneyMTL, a multi-task deep learning framework that uses Multiple Instance Learning (MIL) with task-specific attention pooling to predict severity grades across all five USS attributes simultaneously. Trained on an unprecedented cohort of 50,765 whole-slide images (WSIs), SydneyMTL generates interpretable histologic evidence for clinical practice. In retrospective evaluations against 24 board-certified pathologists, the model achieved an overall mean lenient accuracy of 89.1%, with 22 pathologists exhibiting >80% agreement with the model. When evaluated on an expert-adjudicated “Golden dataset,” the model’s performance improved to 90.2%, demonstrating its capacity to align with multi-expert consensus and filter individual annotator noise. Latent space analysis confirmed that SydneyMTL captures the ordinal structure of the USS, by representing disease severity as a continuous biological spectrum rather than as disjoint categories. Finally, a randomized crossover reader study showed that AI-assisted review significantly reduced interpretation time and improved inter-observer agreement, establishing SydneyMTL as a scalable tool for supporting standardized gastric cancer risk stratification.

**Graphical abstract:** 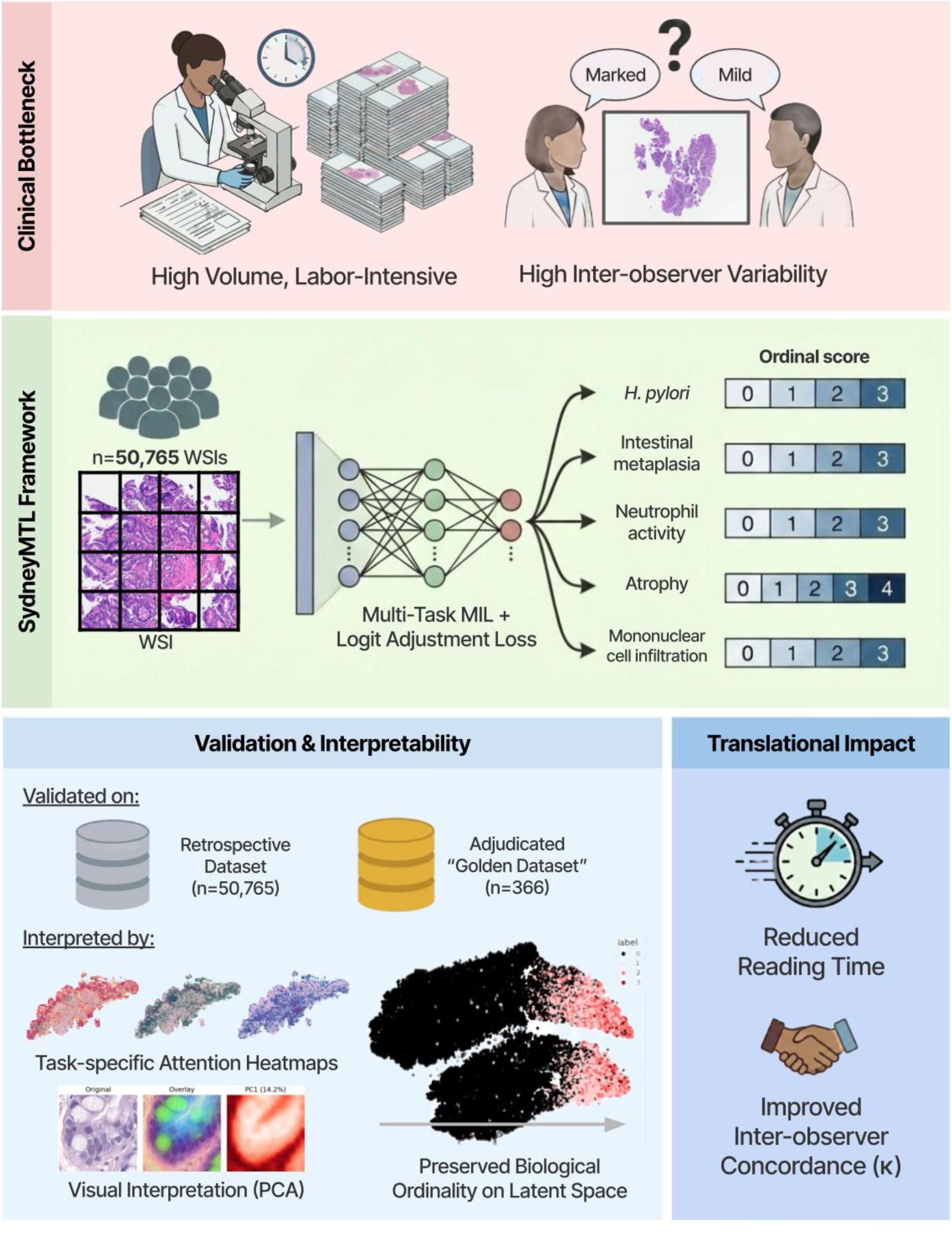

**Highlights:**

- SydneyMTL is the first unified framework to simultaneously predict the full 4-tier severity grades across all five Updated Sydney System attributes.
- Trained on a massive cohort of 50,765 whole slide images, the model aligns with multi-expert consensus on a rigorous “Golden dataset”.
- AI assistance significantly reduces pathologist reading time and harmonizes inter-observer variability in real-world clinical workflows.
- Latent space analysis confirms that SydneyMTL preserves the biological ordinality of disease severity without explicit ordinal constraints.

**The bigger picture:** Gastritis is among the most frequent diagnoses in gastrointestinal pathology, and its histologic severity is central to gastric cancer prevention. In routine practice, pathologists convert subtle mucosal changes into semi-quantitative, ordinal grades using the Updated Sydney System, which evaluates five co-existing histologic dimensions. While this framework provides a shared language, grading is labor intensive and inherently dependent on reader-specific thresholds, creating variability that affects risk stratification and surveillance.

A key concept motivating our study is that gastritis is not defined by a single finding but by multiple criteria that co-occur and interact. This suggests that computational models should learn these criteria jointly – capturing their biological correlations and the continuum of severity – rather than treating each grade as an isolated classification task. SydneyMTL implements this perspective through a unified multi-task, weakly supervised approach that learns directly from a massive cohort of 50,765 routine whole-slide images.

Beyond diagnostic accuracy, our work reveals that the model preserves the ordinality of severity in its representation space, supporting the biological view that discrete clinical categories approximate an underlying continuous biological spectrum. Its attention-based explanations also connect model outputs to interpretable tissue evidence, enhancing clinical trust. Crucially, by harmonizing inter-observer variability, SydneyMTL provides a more reliable foundation for gastric cancer risk assessment, ensuring that premalignant changes are captured with greater consistency. More broadly, our findings reposition AI for gastritis from narrow detection toward scalable, evidence-based decision support that can standardize grading practices and reduce cognitive burden on the global pathology workforce.

## Introduction

Gastritis is a significant global health concern with clinical implications that extend far beyond transient mucosal irritation. Since specific histologic patterns often signal malignant transformation, a thorough evaluation of gastric mucosal injury is essential. The clinical significance of chronic gastritis is most evident in its role as the primary substrate for gastric carcinogenesis. The *Correa Cascade* best articulates this pathological transformation by identifying chronic inflammation as the primary driver of the transition to intestinal-type gastric adenocarcinoma.^1,2^ In this model, *Helicobacter pylori* (*H.pylori*)-induced inflammation drives a stepwise transition from chronic active gastritis to multifocal atrophic gastritis and intestinal metaplasia (IM), establishing a definitive precancerous environment.^3,4^

Accordingly, accurate assessment of gastritis has become a key component of gastric cancer prevention strategies.^5^ Multiple cohort studies have consistently shown that histologic grading of atrophy and IM has clear prognostic value, with higher grades associated with an increased incidence of high-grade neoplasia and invasive cancer.^6,7^ For example, operational staging systems such as OLGA and OLGIM formalize this concept by integrating both the severity and topography of atrophy or IM into stage 0 – IV categories. In this framework, advanced stages (III – IV) reliably identify high risk patients while early stages (0 – II) define a low-risk group.^8^ This risk stratification serves as a foundation for clinical decision-making – informing interventions such as *H. pylori* eradication and endoscopic surveillance – with meta-analyses confirming that such stage-guided management significantly improves risk-adapted outcomes. Consequently, accurate and reproducible assessment of gastritis and its premalignant sequelae is essential to enable robust risk stratification and to support evidence-based treatment decisions.^9,10^

These risk-staging tools are fundamentally derived from histologic attributes defined in the Updated Sydney System (USS), which remains the most widely adopted framework for the pathologic assessment of gastritis.^11^ In practice, OLGA and OLGIM apply the Sydney scores for glandular atrophy and IM across multiple gastric sites to generate cancer risk categories. The USS itself relies on a semi-quantitative 0 – 3 scale encompassing five core histologic features—mononuclear cell infiltration, neutrophil activity, glandular atrophy, IM, and *H. pylori* density—evaluated across a standardized five-site biopsy protocol on the gastric mucosa (two antrum, two corpus, and one incisura). Since OLGA/OLGIM stages directly depend on the accuracy of these underlying Sydney scores, precise and reproducible application of the USS is essential for reliable risk stratification and, ultimately, for effective gastric cancer prevention.^12,13^

Although the USS is the standard framework in routine practice, it is inherently limited by the subjectivity of its semi-quantitative visual scoring. While the USS introduces visual analogue scales to standardize grading, true quantification remains difficult. Multiple studies have documented substantial inter-observer variability among pathologists, with kappa (κ) values for atrophy and neutrophil activity reported as low as 0.04 and 0.22 in routine practice.^11,14^ This inconsistency propagates into risk-stratification systems, introducing uncertainty into gastric cancer risk assessment and surveillance. Furthermore, the USS imposes a significant diagnostic burden; the standardized five-site biopsy protocol requires pathologists to examine multiple tissue fragments across all five semi-quantitative axes, demanding prolonged visual attention and high-fidelity interpretation.

These per-case demands are even more challenging when scaled to real-world clinical volumes. Although the USS recommends five-site gastric sampling, many institutions, especially those in high-throughput or resource-limited settings, commonly submit only one or two biopsy fragments because processing the full protocol is labor-intensive. Even with reduced sampling, laboratories in *H. pylori*-endemic regions handle large volumes of gastric biopsies weekly.^15^ The pressure from this volume, compounded by a global shortage of pathologists, highlights the need for reproducible, scalable computational support. ^16^

Deep learning is transforming digital pathology by enabling expert-level performance in complex diagnostic tasks. ^17–19^ In the context of gastritis, early AI models have primarily focused on narrow objectives, such as detecting only *H. pylori* alone or grading IM and atrophy as independent attributes. ^20^ In real-world gastric biopsies, however, these five USS attributes frequently coexist and are interpreted jointly by pathologists. Consequently, a simultaneous evaluation of these attributes is essential to support accurate risk stratification and informed clinical management.

More recent studies have explored broader approaches. For example, AMMNet used a multi-instance attention framework to predict neutrophil activity, atrophy, and IM based on slide-level labels, achieving high performance across multi-center cohorts.^21^ Similarly, GastritisMIL used semi-supervised learning to detect atrophy and IM and to derive OLGA/OLGIM stages from model outputs.^22^ Although these studies advanced automated gastritis assessment, two fundamental limitations remain. First, no existing framework has comprehensively addressed all five USS attributes. Crucially, *H. pylori*, whose interaction with inflammatory activity is a central driver of chronic gastritis, has been omitted from simultaneous evaluation in prior multi-attribute models. Second, previous approaches often simplify the diagnostic task into binary classifications or truncated scales; to date, no prior study has demonstrated the capability to perform the complete four-tier grading (Absent to Marked) across all five USS attributes simultaneously. Furthermore, these models were trained on relatively modest datasets (1,096 and 2,744 slides, respectively) annotated by a small number of expert pathologists, which restricts the robust clinical generalizability required for diverse real-world pathology.

To address these gaps, we developed a unified, weakly supervised multi-task learning framework that jointly predicts all five Sydney System parameters. We assembled a large-scale cohort of over 50,000 histologically annotated whole slide images (WSIs)—to our knowledge, the largest gastritis dataset to date—capturing the full spectrum of gastric pathology. The model integrates attention-based multiple instance learning with patch-level heatmap visualization to enhance diagnostic transparency. The scale of our dataset enables rigorous evaluation of whether large-scale learning improves reproducibility, strengthens feature-specific consistency, and supports more reliable gastric biopsy interpretation in everyday clinical workflows.

## Results

### Dataset characteristics and inter-observer variability

We analyzed a retrospective cohort of 50,765 gastric biopsy WSIs annotated according to the five histological attributes of the USS (hereafter referred to as the Retrospective dataset). Attributes including *H. pylori*, IM, and neutrophil activity exhibited a pronounced class imbalance characterized by a right-skewed distribution, where prevalence inversely correlated with severity. For instance, *H. pylori* was absent in the vast majority of cases (40,515; 78.81%), while only a marginal fraction was graded as “Marked” (270; 0.53%). In contrast, mononuclear cell infiltration followed a more balanced distribution, centered on “Mild” (63.31%) and “Moderate” (35.20%) grades. Notably, atrophy assessment was precluded in 70.61% of the cohort due to the absence of muscularis mucosae, requiring these cases to be categorized as “Not Applicable (N/A)”.

To counter the prevalence bias of the Retrospective dataset, we constructed a “Golden dataset” (n=366) using a uniform sampling strategy (as described in the Methods). This design ensured adequate representation of higher severity grades and minimized evaluation bias toward the majority “Absent” category. Consequently, the grade distribution was strategically rebalanced: *H. pylori* cases were enriched such that “Moderate” became the most frequent grade (40.98%), and “Marked” IM cases constituted 13.39% of the dataset. This curated composition enables a more stringent and robust evaluation of model performance across the full severity spectrum, particularly for previously underrepresented classes.

Furthermore, an analysis of inter-rater reliability prior to consensus adjudication highlighted the inherent diagnostic complexity of these features. As shown in Table 1, Quadratic Weighted Kappa (QWK) (κ) scores revealed varying levels of agreement among the senior pathologists. While IM demonstrated “substantial” agreement (κ = 0.667), atrophy showed only “fair” concordance (κ = 0.320). These results underscore the challenge of grading subtle or borderline lesions and validate the necessity of our multi-round consensus protocol.^23^

**Table 1.** Diagnostic attributes of the Updated Sydney System in the Retrospective and Golden datasets.

| Cohort | Diagnostic Attributes of USS | Severity (n, (%)) |  |  |  |  | Quadratic Kappa<sup>†</sup> |
| --- | --- | --- | --- | --- | --- | --- | --- |
|  |  | Absent | Mild | Moderate | Marked | N/A |  |
| Retrospective dataset<br>(n=50,765) | *H. pylori* | 40,515 (78.81) | 7,471 (14.72) | 2,509 (4.94) | 270 (0.53) | - | N/A |
|  | Intestinal metaplasia | 34,372 (67.71) | 10,417 (20.52) | 4,967 (9.78) | 1,009 (1.99) | - | N/A |
|  | Atrophy | 10,578 (20.84) | 2,888 (5.69) | 1,093 (2.15) | 359 (0.71) | 35,847 (70.61) | N/A |
|  | Neutrophil activity | 26,080 (51.37) | 12,111 (23.86) | 12,127 (23.89) | 447 (0.88) | - | N/A |
|  | Mononuclear cell infiltration | 18 (0.04) | 32,137 (63.31) | 17,869 (35.20) | 741 (1.46) | - | N/A |
| Golden dataset<br>(n=366) | *H. pylori* | 125 (34.15) | 59 (16.12) | 150 (40.98) | 32 (8.74) | - | 0.556 |
|  | Intestinal metaplasia | 133 (36.34) | 98 (26.78) | 86 (23.5) | 49 (13.39) | - | 0.667 |
|  | Atrophy | 17 (4.64) | 53 (14.48) | 72 (19.67) | 16 (4.37) | 208 (56.83) | 0.320 |
|  | Neutrophil activity | 120 (32.79) | 24 (6.56) | 207 (56.56) | 15 (4.10) | - | 0.543 |
|  | Mononuclear cell infiltration | 0 (0.0) | 84 (22.95) | 252 (68.85) | 30 (8.20) | - | 0.352 |
| †: Inter-pathologist agreement prior to consensus. (Kappa interpretation was calculated using Fleiss' $\kappa$) | | | | | | | |

### Comparative assessment on retrospective and golden cohort

Initial evaluation on the retrospective cohort using 5-fold cross-validation demonstrated high and comparable proficiency across all five USS attributes. As shown in Figure 1, all models closely mirrored the retrospective label distribution and achieved nearly identical performance metrics (e.g. *H. pylori* lenient accuracy >94% across models). While several pairwise comparisons reached statistical significance in lenient accuracy (e.g., atrophy; *p* < 0.05 in lenient accuracy), the absolute improvements were marginal and did not translate into consistently significant differences when evaluated using QWK on the Retrospective dataset. This likely reflects that the models primarily converged to the dominant, systematic annotation patterns present in the large-scale, yet inherently noisy, training labels.

**Figure 1.**
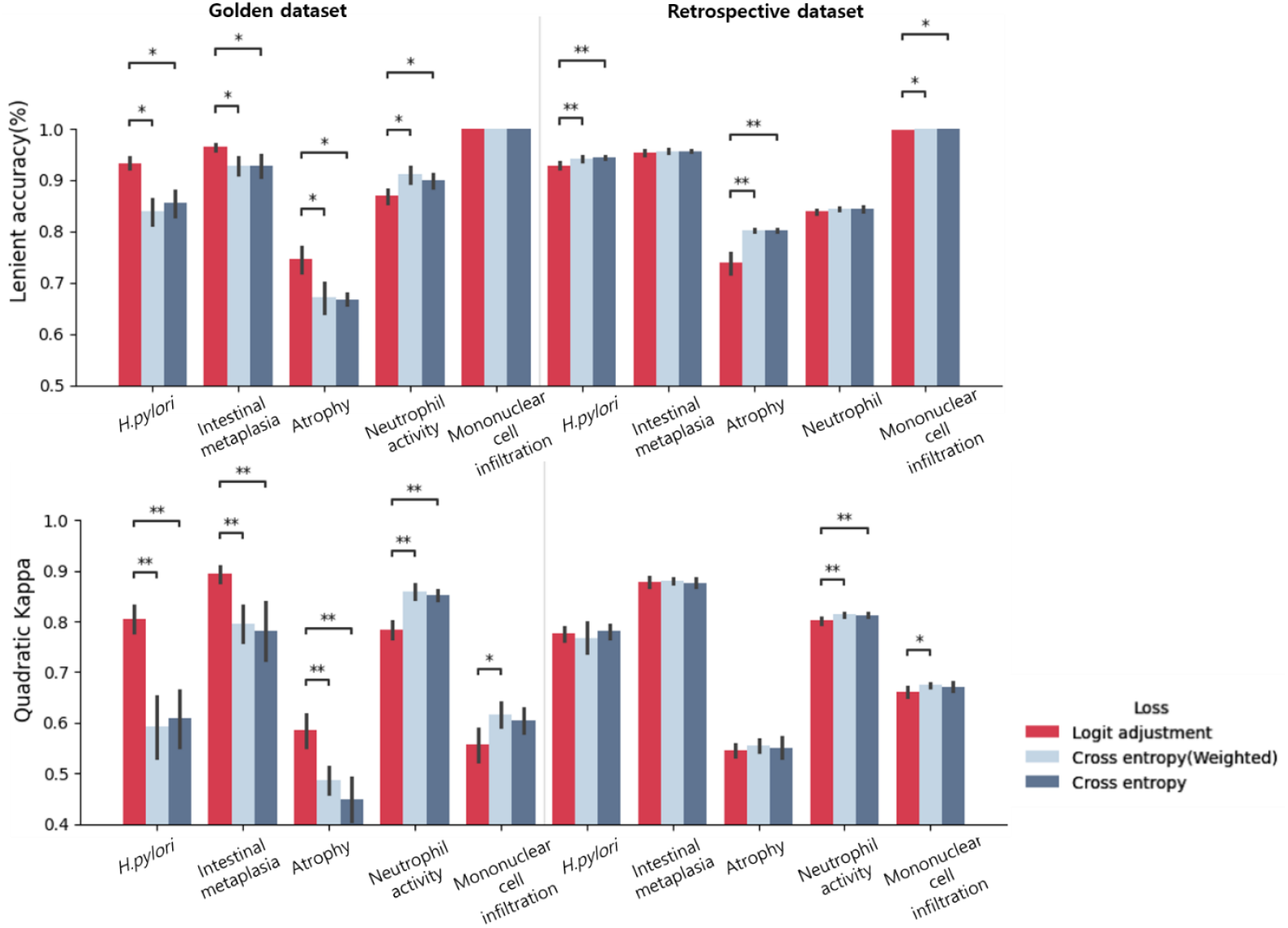
Robustness of SydneyMTL against prevalence-induced bias. While all optimization strategies show high performance on the skewed Retrospective dataset, standard loss functions (Cross-entropy, Weighted Cross-entropy) exhibit a significant performance drop on the balanced Golden dataset. This disparity indicates that standard models are heavily biased toward majority classes, whereas SydneyMTL’s logit adjustment maintains high diagnostic fidelity across the full severity spectrum (See Supplementary figure S1 for class-wise distribution).

Crucially, distinct differences in generalization capability emerged when the models were benchmarked against the Golden dataset. Unlike the baseline frameworks, which exhibited a marked drop in performance when transitioning to the adjudicated standard, SydneyMTL maintained substantially higher agreement and accuracy. It simultaneously preserved superior calibration across the full severity spectrum rather than collapsing its predictions toward the skewed majority grades. Specifically, for *H. pylori* grading, the baseline QWK score plummeted to 0.592, whereas SydneyMTL achieved a robust score of 0.826 (*p* < 0.01), with lenient accuracy reaching 95.1%. Similarly, for IM, our framework improved the QWK score from 0.796 to 0.898 (*p <* 0.05), significantly outperforming standard loss functions.

Across all metrics, SydneyMTL demonstrated consistent superiority on the golden standard, with notable gains observed in both lenient accuracy and QWK across multiple USS attributes. Most remarkably, SydneyMTL outperformed the baseline by over 0.1 QWK points (0.599 vs. 0.487) when evaluating glandular atrophy—a feature notoriously characterized by high inter-observer variability. These results confirm that our proposed uncertainty-weighted strategy successfully encourages the model to learn invariant morphological features and prioritizes objective diagnostic signals over the dominant, biased patterns of the training set.

### Robustness of the model across 24 individual pathologists

To assess the clinical robustness of our AI model, we evaluated its performance on the Retrospective dataset by comparing its predictions against the independent diagnoses of 24 individual pathologists. Overall, SydneyMTL demonstrated strong alignment with the majority of human readers. The model achieved over 80% average lenient accuracy with 21 out of the 24 pathologists and exceeded 90% agreement with a subset of four pathologists (Figure 2). Among the five USS attributes, the model exhibited particularly robust performance for IM and *H. pylori*. The average lenient accuracy for IM was 93.2%. Similarly, for *H. pylori*, the model achieved an average accuracy of 93.9%, with even the lowest observed agreement remaining relatively high at 84.6%, indicating stable and reliable diagnostic capability across different observers.

**Figure 2.**
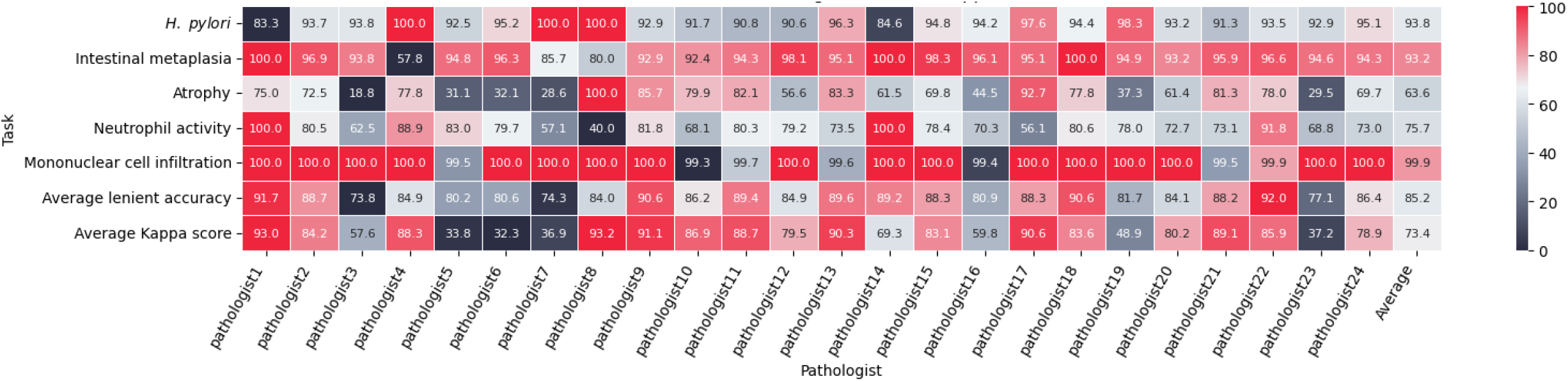
Diagnostic performance heatmap of 24 pathologists on the Golden dataset. The heatmap displays the lenient accuracy for each of the five Updated Sydney System attributes (rows) across 24 individual pathologists (columns). To visualize the relative performance distribution within each task, the color intensity is normalized row-wise.

In contrast, the assessment of glandular atrophy revealed significant variability in model-pathologist agreement. While the average lenient accuracy for atrophy was 63.6%, performance fluctuated drastically across individuals, dropping to as low as 18.8% against Pathologist 3. This marked heterogeneity aligns with the high inter-observer variability well-documented in human atrophy assessment, suggesting that the model encounters the same “borderline” diagnostic challenges as human experts. Regarding mononuclear cell infiltration, the model achieved near-perfect lenient accuracy (approaching 100%). This high level of concordance was expected, given the class distribution where the vast majority of cases were graded as mild or moderate (Grade 1 or 2). Finally, the overall robustness of the model was further validated by an average QWK (κ) score of 0.734 across all 24 pathologists, confirming its high utility as a stable diagnostic aid.

### AI impact on observer variability and interpretation time

The inherent subjectivity in quantifying chronic gastritis, particularly in borderline cases, necessitates an objective assessment of whether AI assistance can harmonize diagnostic thresholds among pathologists. Our crossover study revealed that the introduction of SydneyMTL significantly mitigated inter-observer variability, which is a key challenge in the reproducible application of the Sydney System. In the unassisted review phase, the mean QWK (κ) between the two senior pathologists stood at κ = 0.751 ± 0.016. However, when pathologists utilized the real-time AI guidance, this concordance improved to κ = 0.774 ± 0.02. A granular analysis showed that this convergence was most pronounced in high-variance tasks such as Mononuclear cell infiltration, which improved from κ = 0.454 to 0.580. This convergence indicates that the model functions as an objective digital reference point, effectively guiding experts to align their subjective interpretations of lesion severity and extent, thereby enhancing the standardization of the Sydney grading system.

In addition to improving reproducibility, AI integration substantially enhanced diagnostic efficiency (Figure 3). We measured the time required for the holistic assessment of the five attributes per WSI. The average interpretation time per WSI decreased by 34.3% with the assistance of SydneyMTL (linear mixed-effects model; the coefficient for the AI-assisted condition (vs. unassisted) was significantly negative, *p* <.001). Specifically, Pathologist 1 achieved a 38.7% reduction (paired Cohen’s *d* = 0.73), while Pathologist 2 achieved a 29.7% reduction (paired Cohen’s *d* = 0.65). Since pathologists perform a simultaneous, integrated assessment of multiple features, this overall time reduction confirms that the model’s interpretable attention maps—which rapidly highlight key regions such as *H. pylori* clusters or focal IM—effectively reduce the cognitive load and search time required to reach a final diagnostic conclusion.

**Figure 3.**
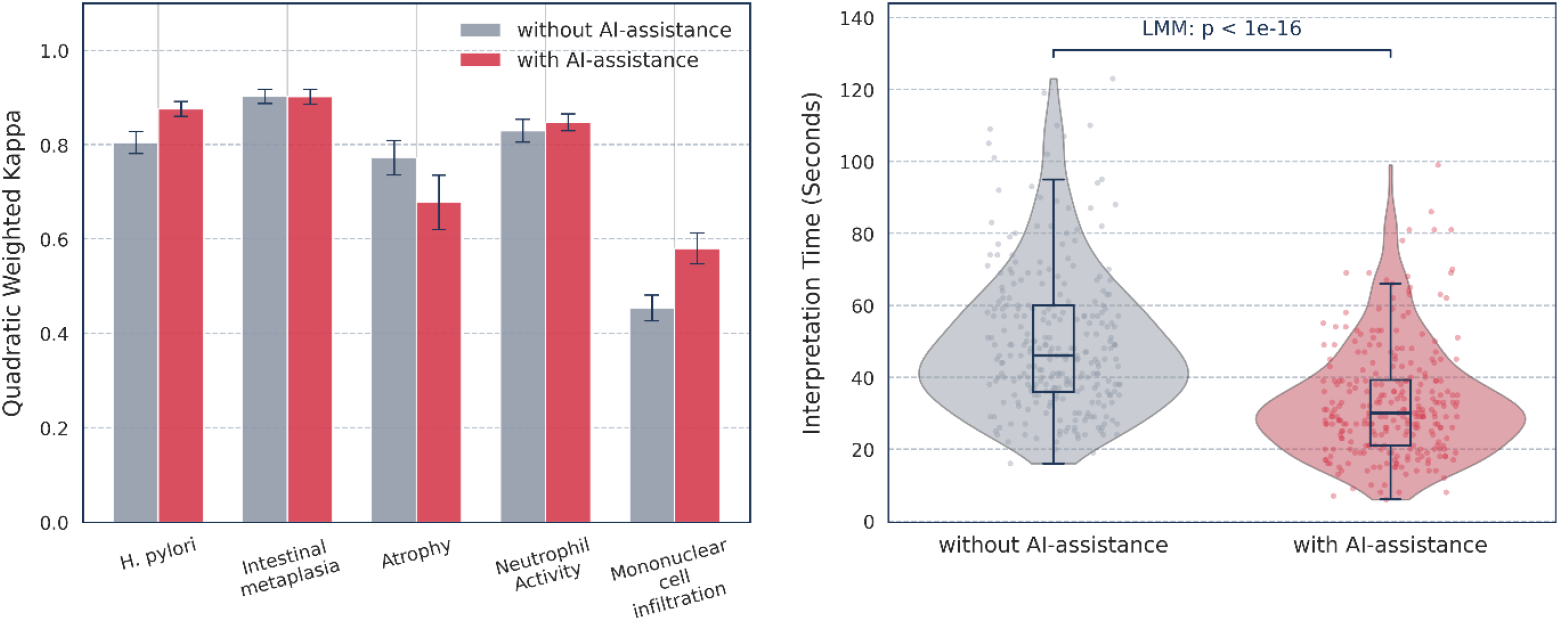
Impact of SydneyMTL on diagnostic harmonization and clinical workflow efficiency. (Left) Inter-observer agreement across five attributes. AI assistance significantly mitigated variability by providing an objective digital reference, leading to a notable convergence in high-variance tasks such as mononuclear cell infiltration. (Right) Enhancement of diagnostic efficiency. SydneyMTL integration achieved a 34.3% overall reduction in interpretation time (*p* < 0.001), effectively lowering cognitive load and search time for both pathologists.

### Preservation of biological ordinality in latent feature space

To validate that the model’s representational logic reflects the biological progression of gastric pathology, we examined the topological structure of its latent feature space. Specifically, we assessed whether the model captures the intrinsic ordinal nature of the USS grades, which is a critical requirement for clinical plausibility and diagnostic reliability.

Visualizing the slide-level embeddings of the Golden dataset via t-distributed Stochastic Neighbor Embedding (t-SNE) revealed a highly structured and topologically coherent manifold (Figure 4). Key attributes, including *H. pylori* density, neutrophil activity, and IM, exhibited a sequential trajectory corresponding to severity gradients (transitioning from “Absent” to “Marked”) rather than forming disjoint, isolated islands. This spatial arrangement suggests that the model successfully encodes the semantic continuity of disease progression within its latent representations.

**Figure 4.**
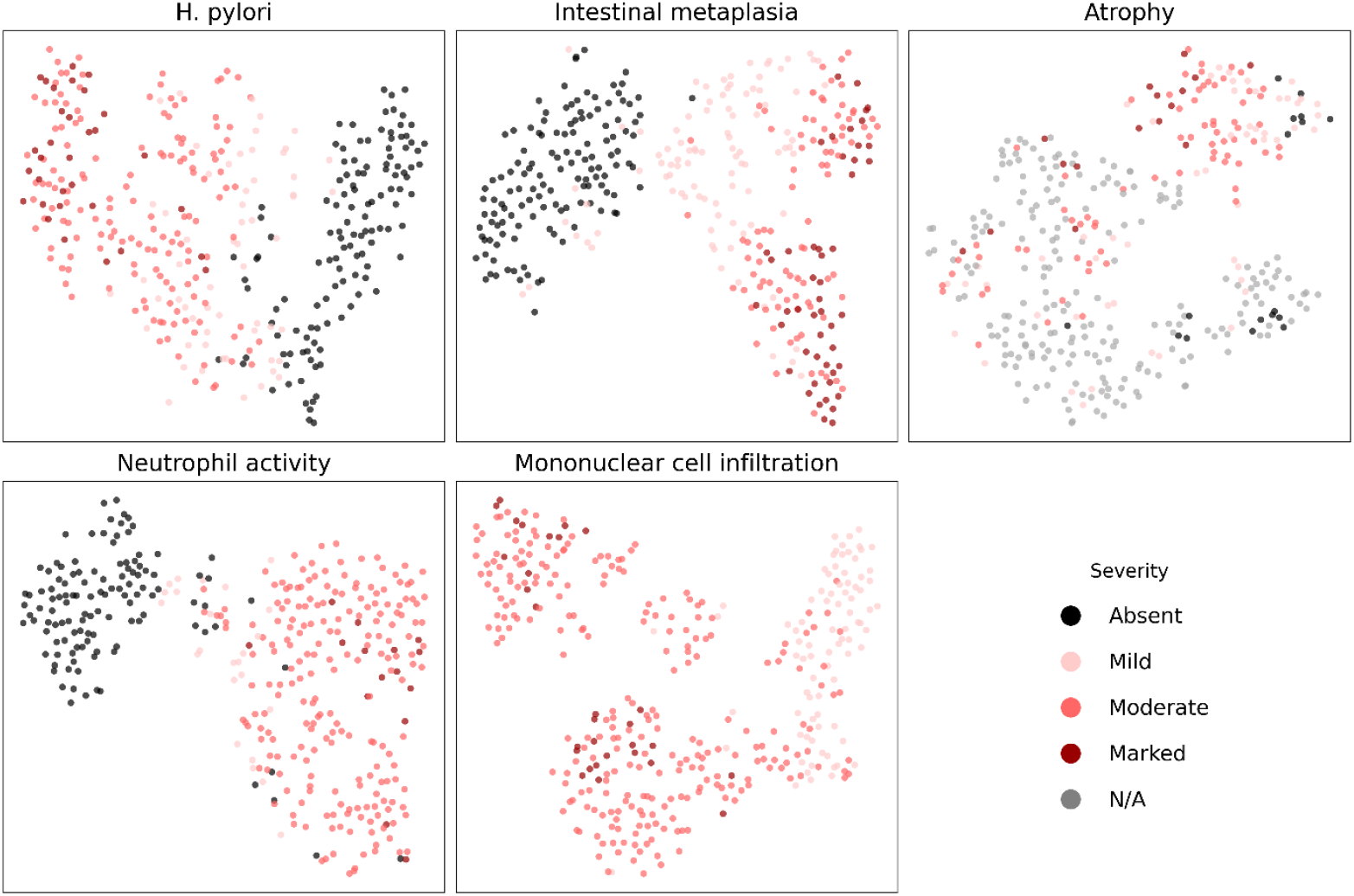
t-SNE visualization of slide-level embeddings reveals a structured manifold. We mapped the high-dimensional embeddings of the Golden dataset into a two-dimensional space, revealing that the model captures the biological continuity of gastritis. Rather than forming isolated clusters, key histological attributes exhibit a sequential trajectory that mirrors clinical severity (from “Absent” to “Marked”).

We quantitatively substantiated these visual patterns by calculating the interclass Euclidean distances between embedding centroids (Supplementary figure S2). This analysis revealed a strict monotonic relationship within the feature space; distances were minimized between adjacent severity grades and maximized between divergent ones. For *H. pylori*, the Euclidean distance from the “Absent” centroid increased in a near linear-fashion relative to severity: 5.88 for “Mild”, 9.15 for “Moderate”, and 11.90 for “Marked”. This geometric evidence confirms that the model does not treat diagnostic categories as independent entities; instead, it has successfully learned the underlying biological spectrum of gastritis severity, ensuring that its latent feature space reflects clinical reality.

### Ablation test – optimization of logit adjustment temperature

While logit adjustment improved performance across all tasks, the temperature parameter (τ) necessitated a strategic trade-off. We strategically optimized the logit adjustment temperature to prioritize diagnostic sensitivity for the “premalignant triad” (*H. pylori* density, IM, and atrophy). While a lower temperature (τ = 0.5) yielded the highest global average lenient accuracy (0.900), τ = 0.75 ensured superior performance on the clinically consequential subset (0.876 vs. 0.870 at τ = 0.5) (Table 2). By deliberately choosing this operating point, we align SydneyMTL’s predictive focus with the high-stakes features essential for gastric cancer risk stratification systems, such as OLGA/OLGIM, rather than merely maximizing raw global performance (Table 2).

**Table 2.** Ablation test of Logit Adjustment temperature (τ) for clinical performance optimization.

| $\tau$ | Lenient accuracy on Golden dataset (5-fold average, [SD]) | | | | | Average | Premalignant Triad* |
| --- | --- | --- | --- | --- | --- | --- | --- |
|  | *H. pylori* | Intestinal metaplasia | Atrophy | Neutrophil activity | Mononuclear cell infiltration |  |  |
| **0.5** | 0.913<br>[0.015] | 0.953<br>[0.014] | 0.745<br>[0.008] | 0.889<br>[0.01] | 1.000<br>[0.01] | 0.900 | 0.870 |
| **0.75** | 0.928<br>[0.007] | 0.960<br>[0.011] | 0.74<br>[0.032] | 0.863<br>[0.019] | 0.997<br>[0.004] | 0.898 | **0.876** |
| **1** | 0.913<br>[0.015] | 0.959<br>[0.01] | 0.651<br>[0.016] | 0.833<br>[0.02] | 0.793<br>[0.052] | 0.830 | 0.840 |

\* Premalignant Triad – *H. pylori* density, Intestinal metaplasia, and Atrophy

### Visual interpretation

We analyzed the spatial distribution of attention weights to verify whether the model’s decision-making process aligns with clinically established diagnostic regions. For *H. pylori*, attention heatmaps were predominantly concentrated on the superficial epithelium (the outer layer of glands) (Figure 5-Left). Notably, the patches with the highest attention weights directly captured distinct colonies of *H. pylori* (Supplementary figure S3). Similarly, in cases of IM, the model consistently assigned high attention scores to glandular structures containing characteristic goblet cells. For inflammatory activity, the heatmaps for neutrophils specifically highlighted glands exhibiting intra-gland infiltration. For mononuclear cell infiltration, the model focused on the regions characterized by a high density of lymphocytes infiltration. In cases of marked atrophy, the model’s attention was preferentially localized to the muscularis mucosae and adjacent gland bases – an anatomical prerequisite for grading glandular atrophy in the USS. Notably, in cases lacking the muscularis mucosae where assessment was precluded, the model appropriately reflected this limitation by categorizing them as “N/A”.

**Figure 5.**
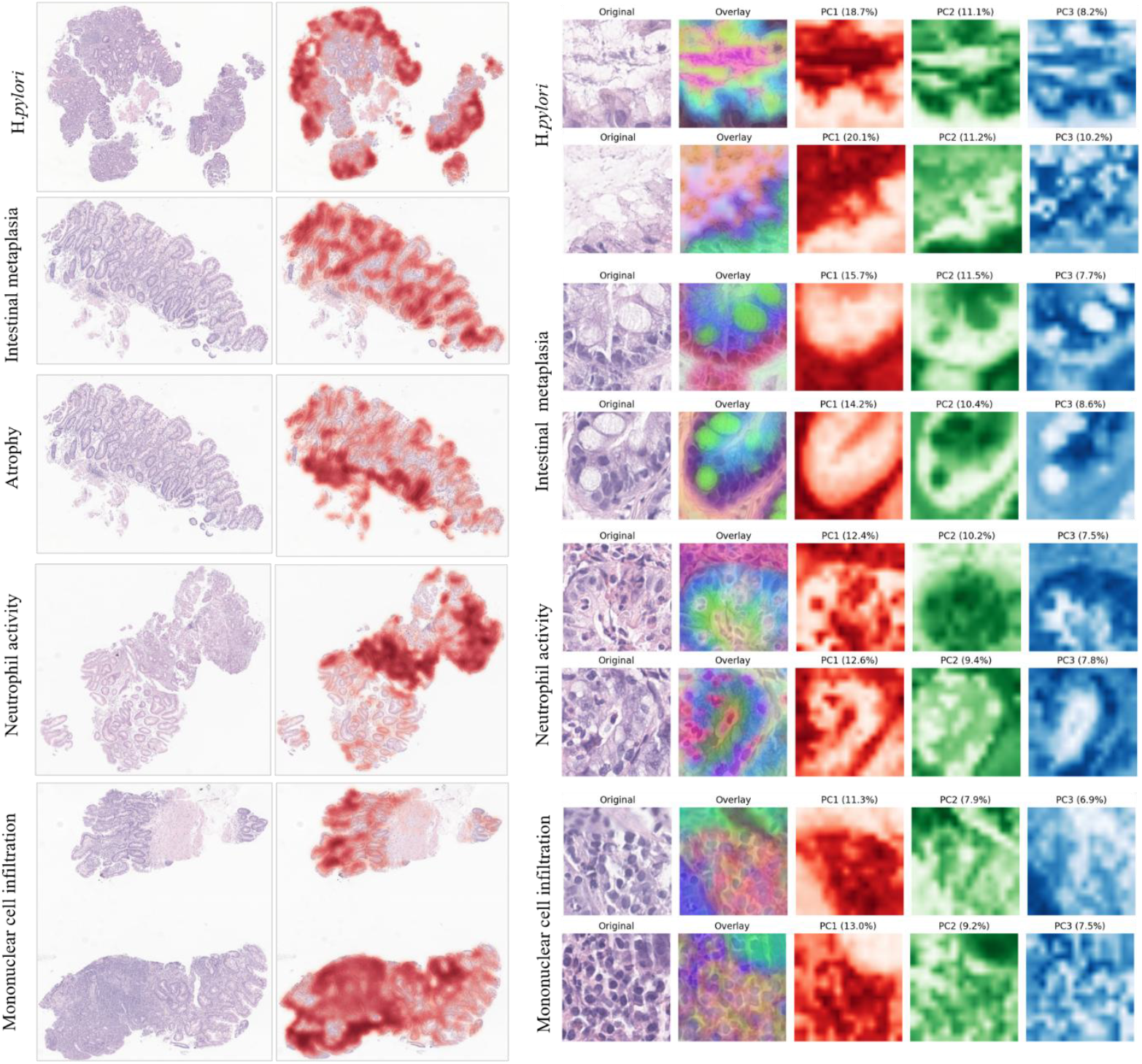
Multi-scale visualization of diagnostic interpretability and feature representations. (Left) Slide-level interpretation using task-specific attention weights. The heatmaps illustrate the model’s ability to localize diagnostic regions corresponding to each of the five USS attributes across the whole slide image. (Right) Visualization of learned feature representations using Principal Component Analysis (PCA). Spatial activation maps of the top three principal components (PC1, PC2, PC3) are derived from patch-level embeddings for *H. pylori*, neutrophil activity, intestinal metaplasia, and mononuclear cell infiltration. These maps demonstrate the model’s capacity to disentangle specific diagnostic features, such as bacterial aggregates and goblet cells, from the tissue background. Glandular atrophy is excluded from this patch-level PCA visualization as its pathology is primarily defined by the structural depletion or loss of glandular units rather than the presence of discrete cellular objects, making architectural context more informative than individual patch-level representations.

Complementing the slide-level attention analysis, we further investigated whether the model captures specific microorganisms and morphological features at the patch level. To verify this, we applied Principal Component Analysis (PCA) to the patch embeddings to visualize the dominant features learned by the model. For slide with “Marked” *H. pylori* severity, the activation maps of the first two principal components (PC1 and PC2) were strongly localized to small regions corresponding to the bacterial aggregates (Figure 5-Right). Notably, these components specifically highlighted characteristic aggregates of *H. pylori* within the surface mucus and gastric pits while effectively suppressing the background tissue and artifacts (Figure 5-Right).

For IM, the second principal component (PC2) showed a high correlation with goblet cells, the hallmark feature of the condition, selectively activating mucin-filled regions. Regarding inflammatory cells, the model successfully disentangled distinct morphological subtypes; for neutrophil activity, both PC1 and PC2 concentrated on segmented nuclei with multiple lobes, whereas for mononuclear cells, PC1 specifically highlighted small patches containing mononuclear cells. These results demonstrate that the model constructs its predictions based on biologically relevant morphological evidence rather than confounding background signals.

## Discussion

Accurate histological grading of gastric biopsies is essential for gastric cancer prevention and serves as the basis for risk stratification systems. However, the routine application of the USS is currently hindered by substantial inter-observer variability and the labor intensive nature of multi-feature assessment. In this study, we introduce SydneyMTL, the first unified framework capable of simultaneously performing four-tier grading for all five Sydney System attributes. Developed and rigorously validated on the largest cohort of gastric biopsies to date (n = 50,765) and adjudicated dataset, our model captures the comprehensive histological landscape of chronic gastritis. By automating the grading process with strict adherence to biological ordinality and expert consensus, this work represents a paradigm shift from subjective visual estimation to reproducible, quantitative computational pathology.

The primary significance of SydneyMTL lies in its ability to overcome the “subjectivity bottleneck” inherent in human grading. Previous deep learning attempts have typically been constrained by small datasets (< 3,000 images) derived from few observers. This limits generalization and increases the risk of overfitting to specific human biases. In contrast, our framework uses a large training set of 50,765 WSIs annotated by 24 board-certified pathologists. The model’s ability to achieve consistent agreement across 21 of these 24 pathologists demonstrates a robust, unbiased predictive capability. Rather than mimicking individualized diagnostic tendencies, the model learns invariant features that generalize across the broad spectrum of diagnostic patterns found in routine practice. Building on this diverse training foundation, we validated the model against a “Golden dataset” established through multi-round expert consensus. This demonstrated that SydneyMTL effectively harmonizes individual diagnostic variations to align with a multi-expert consensus standard. Furthermore, our use of Logit Adjustment loss specifically addresses the extreme class imbalance typical of screening population. As evidenced by the significant gains in recall for high-severity grades in Supplementary figure S1, this approach encourages the model to confidently identify rare but critical “Marked” lesions—such as dense *H. pylori*—which are frequently under-predicted by standard algorithms biased toward the majority class. By effectively recalibrating the decision boundaries, SydneyMTL ensures that high-risk cases are not obscured by the prevalence of mild or normal samples.

Beyond diagnostic metrics, SydneyMTL is defined by its biological transparency and interpretability. Our manifold analysis (t-SNE) and principal component analysis (PCA) confirm that the model bases its decisions on established morphological evidence rather than confounding artifacts. Specifically, the model has learned to explicitly map the bacterial aggregates of *H. pylori* and the mucin-filled goblet cells of IM to specific latent features, effectively separating them from background noise. Importantly, the attention mechanism exhibited spatial logic consistent with gastric anatomy. It correctly localized *H. pylori* colonization to the luminal surface epithelium while mapping mononuclear inflammation to the lamina propria. For glandular atrophy, the attention maps concentrated along the muscularis mucosae and the adjacent gland bases, reflecting the anatomical compartment where gland loss and basal gland remodeling are assessed (Figure 5). Consistent with the Sydney System workflow, the model recognized that meaningful atrophy grading requires the presence of muscularis mucosae; cases lacking this landmark were appropriately categorized as N/A, validating that the model respects the topographical constraints of tissue architecture. Furthermore, the model’s latent architecture autonomously reorganizes slide-level embeddings into a structured manifold that aligns with the USS’s ordinal hierarchy. Remarkably, the model learned to place “Moderate” cases between “Mild” and “Marked” grades without explicit ordinal constraints during training. This emergent preservation of ordinality indicates that SydneyMTL captures the continuous spectrum of gastritis severity, thereby treating disease progression as a continuous biological spectrum rather than as a series of disjoint, independent categories.

Despite the standardization goals of the USS, histologic grading remains challenging due to subjective decision thresholds, leading to only fair-to-moderate inter-observer agreement (κ~0.31 for atrophy and 0.62 for IM in prior studies).^24^ Consistent with this literature, our baseline evaluation showed the lowest concordance in glandular atrophy, likely reflecting the inherent difficulty in operationalizing the phenotype for gland loss and mucosal architecture (κ=0.320). To bridge this gap, our framework was trained to internalize a broader consensus of expert decision-making. Notably, in a crossover study involving two board-certified pathologists, AI-assisted evaluation significantly improved inter-observer concordance for high-variance tasks; for instance, the QWK for mononuclear cell infiltration rose from 0.454 to 0.580, while *H. pylori* detection reached near-perfect alignment (κ = 0.876). The resulting high performance suggests that the model can serve as a consistent objective reference, successfully navigating the subtle diagnostic boundaries that typically trigger discordance among human readers.

Crucially, when pathologists performed AI-assisted interpretation with SydneyMTL, the overall mean inter-observer agreement improved significantly (mean QWK from 0.751 to 0.774). This gain was observed even when readers were free to override the model, suggesting the model’s initial recommendations provided a highly credible “clinical baseline.” Furthermore, mean review time decreased by 34.2% (Mean: 50.6 sec vs. 33.3 sec; *p* < 0.001), marking a significant departure from previous findings. For instance, prior research on Giemsa-stained gastric biopsies using CNNs reported that AI assistance actually increased review time, potentially because pathologists performed more exhaustive manual verification due to uncertainty in model reliability. In contrast, the efficiency gains in our study—characterized by a large effect size (Cohen’s *d* of 1.00 for P1 and 0.83 for P2)— indicate that SydneyMTL’s outputs were sufficiently robust for readers to incorporate them without triggering a ‘confirmatory search’ burden, thereby streamlining the digital pathology workflow.

Our study has several limitations. First, despite the large-scale cohort, the data were derived from a single high-volume center. Nevertheless, while prior studies typically relied on a limited number of pathologists for evaluation, our study benefits from a more generalizable framework by involving 24 diverse annotators within a single center. To further ensure generalizability across different diagnostic cultures, external multi-institutional validation is warranted in future work. Second, while our multi-task formulation improved overall robustness, multi-objective optimization can occasionally induce task interference. Future iterations will explore gradient-balancing approaches (e.g., GradNorm) to further refine per-attribute performance. Finally, as this evaluation remains retrospective, we plan to integrate SydneyMTL into the laboratory information system for prospective studies to quantify its real-world impact on diagnostic efficiency and the reproducibility of gastric cancer risk stratification.

In conclusion, SydneyMTL represents a paradigm shift in gastric pathology as the first unified framework to simultaneously assess all five attributes of the USS. By leveraging an unprecedented large scale dataset and adjudicated dataset, the model transcends individual observer bias and aligns with a rigorous multi-expert consensus. The demonstrated reduction in interpretation time and the harmonization of inter-observer variability underscore its potential as a scalable decision-support tool in high-volume clinical workflows. SydneyMTL promises to enable standardized, reproducible gastritis grading in routine practice, supporting more timely and accurate gastric cancer risk stratification globally.

## Method

### Data collection and annotation

We assembled a large dataset of gastric biopsy specimens from Seegene Medical Foundation (SMF), a high volume reference diagnostic pathology laboratory in South Korea. The specimens were obtained between February 2024 and July 2025. They were processed using routine histopathological protocols, including formalin fixation, paraffin embedding, 3μm microtome sectioning, and hematoxylin and eosin (H&E) staining. All slides were digitized at 40x magnification (0.25 μm/pixel) using a Leica Aperio GT450 scanner at SMF.

Board-certified pathologists at SMF reviewed all H&E-stained slides and assigned labels according to the USS. The USS defines five key histologic attributes: mononuclear cell infiltration, neutrophilic activity, glandular atrophy, IM, and *H. pylori* infection. Each attribute is graded on a four-point ordinal scale (0 = Absent, 1 = Mild, 2 = Moderate, 3 = Marked). To ensure high clinical fidelity, we integrated “N/A” (Not Applicable) as a distinct category for glandular atrophy, addressing cases where the absence of the muscularis mucosae precludes definitive grading. By incorporating the full spectrum of labels including “N/A”, SydneyMTL better reflects real-world diagnostic variability and increases robustness to partially evaluable specimens (Table 3). Unlike previous studies that exclude such cases, our framework explicitly learns these evaluation constraints, mimicking the high-fidelity decision-making process of pathologists who must identify non-evaluable tissue in routine biopsy scenarios.

**Table 3.** Summary of Histological Grading Criteria (Updated Sydney System)

| Histologic attributes of the USS | Description | Grade |
| --- | --- | --- |
| *H. pylori* | Graded based on the density of *Helicobacter pylori* colonization on the surface mucus and gastric pits. | 0-3 |
| Intestinal metaplasia | Defined as the replacement of the gastric mucosa by intestinal-type epithelium, characterized by the presence of goblet cells. | 0-3 |
| Glandular atrophy | Evaluated by the loss of inherent gastric glands. | 0-3, N/A |
| Neutrophilic activity | Graded according to the presence of polymorphonuclear leukocytes (neutrophils) within the surface epithelium or glandular lumens. | 0-3 |
| Mononuclear cell infiltration | Graded based on the density and spatial distribution of mononuclear cell infiltration within the lamina propria. | 0-3 |

In addition, to ensure a rigorous and unbiased evaluation, we created an independent “Golden dataset” (n=366), which serves as a high-quality reference standard. The development dataset (training/internal validation) comprised slides collected between February 2024 and July 2025, while the Golden dataset was assembled from a strictly non-overlapping period (August–September of 2025) to guarantee complete temporal separation. For establishing ground truth, three senior pathologists with over 15 years of experience independently reviewed all cases. To resolve inter-observer discordance and ensure label reliability, the final ground truths were determined through a multi-round consensus adjudication process. Importantly, this held-out set was intentionally curated to offer a more balanced representation than the naturally skewed distribution of clinical practice. To ensure a rigorous evaluation across the full diagnostic spectrum, we focused on the relationship between *H. pylori* and IM, carefully selecting samples to include a wide variety of their severity combinations. By prioritizing the inclusion of rare clinical presentations and moderating the volume of more common, mild cases, we established a “Golden dataset” where each diagnostic grade—from absent to marked—is adequately represented. While the remaining three attributes were sampled alongside these primary markers, this comprehensive selection ensures that our performance metrics reflect the model’s true capability across all stages of gastritis, preventing the results from being disproportionately influenced by the high prevalence of “Absent” cases.

### Problem definition

In routine gastric biopsy interpretation, pathologists assign five USS histologic attributes grades simultaneously on the same WSI: mononuclear cell infiltration, neutrophil activity, atrophy, IM, and *H. pylori* density. Each feature is graded into four ordinal categories (e.g., 0–3, except for atrophy). Therefore, for a single specimen, the diagnostic target is not a single label but a 5-dimensional grade vector predicted concurrently from one WSI. Formally, Let the set of tasks be 𝒯, where each *t* ∈ 𝒯 corresponding to one Sydney attribute. For slide *i*, only slide-level labels are available (weak supervision): 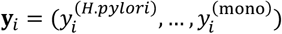, where 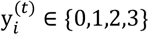 (for atrophy, 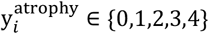 where 4 refers to “N/A”). Given a WSI *X*_*i*_, our goal is to jointly learn a multi-task predictor that outputs task-specific grade distribution for all five USS attributes. Formally, for each task *t* ∈ 𝒯, the model predicts a logit vector 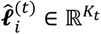, where *K*_*t*_ *= 4* for *t ≠ atrophy* and *K_t_ = 5* for atrophy (with the additional class corresponding to N/A). The parameters *θ* are learned by minimizing a multi-task objective function over the dataset 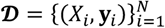: 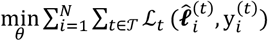, where ℒ_*t*_represents the task-specific loss.

### Unified multi-Task MIL framework for Sydney grading

To simultaneously predict all Sydney attributes, we adopted a hard parameter sharing multitask learning (MTL) framework integrated with multiple instance learning (MIL) (Figure 6). Each WSI *X*_*i*_ is decomposed into *M*_*i*_ fixed-size patches to form a bag *X*_*i*_ *=* {*x*_*i,1*_, *…, x*_*i,Mi*_}. A feature extractor extracts patch embeddings ***h***_*i,j*_ = *f*(*x*_*i,j*_) ∈ ℝ^*d*^. These patch embeddings are first passed through a shared adaptor network *g*_*ϕ*_ to project them in to task-agnostic latent space: ***z***_*i,j*_ *= g*_*ϕ*_(***h***_*i,j*_). To model Sydney attribute-specific evidence, we employ task-specific attention pooling for each task *t*. Given the set of projected embedding 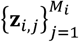, and attention module 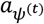 produces a normalized attention weights for each patch 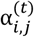. A task-specific slide-level vector is then obtained via attention-weighted aggregation: 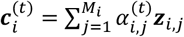. The independent attention modules identify regions most relevant to each histologic criterion. The task-specific classification heads map these contextual representations to the Sydney grades. Finally, each task has an intendent linear classification head 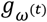 that maps 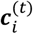 to logits over grade categories: 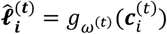. Then probability vector 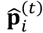 was generated with softmax function, and final predictive label 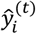 was calculated with argmax function. All components are trained end-to-end with weak slide-level supervision by minimizing the sum of task-wise Cross-entropy losses.

**Figure 6.**
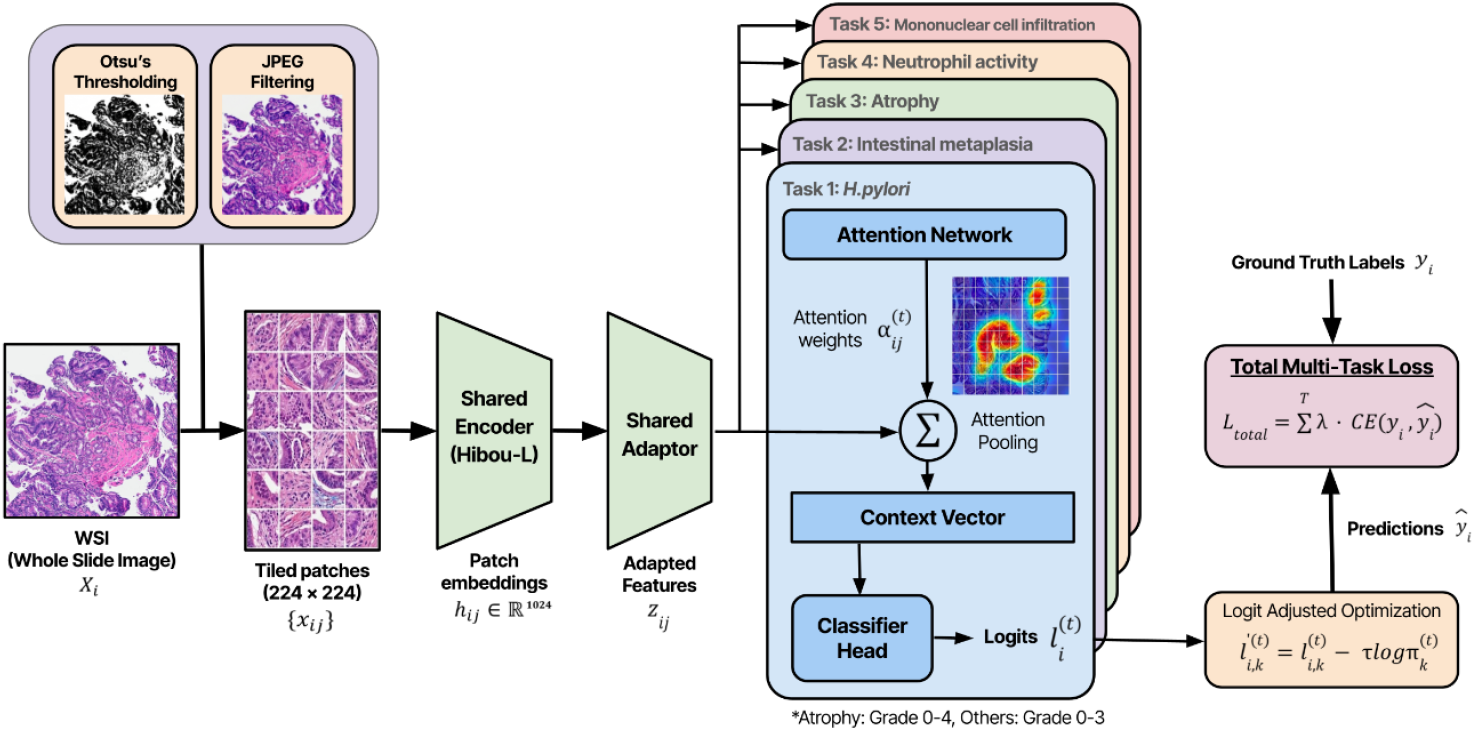
Overview of the unified multi-task MIL framework for Sydney system grading

### Prior-based logit adjustment for Long-Tail Sydney grading

The Sydney System grade distribution in routine gastric biopsy datasets is inherently imbalanced. Most specimens exhibit absent or mild findings, while moderate or severe grades constitute only a small fraction of cases. This long-tailed pattern was present across all five diagnostic tasks in our dataset of 50,765 slides. For instance, “Marked” grades appeared in fewer than 2% of slides, depending on the Sydney attributes, whereas “Absent” grade constituted the majority. These distributions reflect actual clinical prevalence rather than dataset bias; however, they present significant challenges for optimization under standard Cross-entropy training.

To better accommodate this imbalance, we employed Logit Adjustment, a statistically grounded method for long-tail learning introduced.^26^ Logit adjustment incorporates empirical class priors into the model’s logits, encouraging more appropriate decision margins between common and rare labels.

For each Sydney task, we compute class priors as: 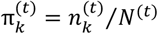 where 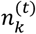 denotes the number of training slides assigned to grade *k* for task *t*, and 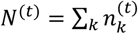 During training, we adjust the logit the task-specific logits 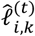 by incorporating the log-prior: 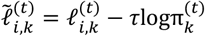 where *τ* is a temperature hyperparameter controlling the strength of the adjustment. The adjusted logits 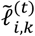 are then used to compute the Cross-entropy loss for each task, and the overall objective is obtained by summing losses across tasks. This formulation is Fisher-consistent with respect to the balanced error rate and avoid limitations associated with traditional reweighting or margin-based losses under class imbalance.^26^ The procedure was applied independently to all five Sydney tasks (including the extended five-class formulation for atrophy). Through systematic empirical calibration, we found that temperature values between 0.5 and 0.75 yielded stable training and improved the recognition of infrequent high-grade categories, while maintaining robust optimization within the multitask MIL framework.

### Statistical methods

To comprehensively evaluate SydneyMTL, we designed a multifaceted experimental framework focusing on predictive performance, unbiased concordance across multiple pathologists, clinical efficacy, preservation of ordinality, and semantic interpretability. The detailed protocols are as follows:

1. **Model performance metrics**: We quantified model performance using both stratified five-fold cross-validation and Golden dataset to account for the significant class imbalance in Sydney grade distributions. Given the ordinal and semi-quantitative nature of the USS, we reported two primary metrics:
  - **Quadratic Weighted Kappa**^27^: This serves as a robust metric for ordinal data, reflecting the graded severity of misclassification by imposing greater penalties for larger disagreements.
  - **Clinically Lenient Accuracy**: To better align model evaluation with real-world practice, we prioritized this metric to account for inherent inter-observer variability in subjective thresholds. Lenient accuracy incorporates a one-grade tolerance for pathological severities (Grade 1-3) while maintaining a strict zero-tolerance policy for the boundary between normal tissue (Grade 0) and pathological change (Grade *≥*1).^28^ A prediction ŷ is considered correct relative to the ground truth *y* if:
  1 For *y = 0*: The prediction must be ŷ *= 0*.
  2 For *y ≥ 1*: The prediction must satisfy *ŷ ≥ 1* and | *ŷ ™ y*| *≤ 1*.
2. **Unbiased concordance analysis**: To verify that the model generalizes beyond a single annotator, we evaluated concordance on a test dataset labeled by multiple pathologists in the Retrospective dataset.
3. **Clinical efficacy (Crossover study)**: To evaluate the model’s practical utility in real-world diagnostic workflows, we employed a randomized crossover design. The study involved senior pathologists reviewing slides in two phases separated by a 5-week washout period. To eliminate memory effects, the sequence of reviews was randomized: one group of readers began with the AI-assisted review, while the other started with the unassisted view. We conducted a randomized crossover reader study using a dedicated cohort of 134 WSIs. This experiment aimed to quantify the value of SydneyMTL as a decision-support tool that enhances human performance, rather than merely acting as a standalone classifier. Diagnostic performance was assessed using lenient accuracy and QWK. Workflow efficiency was evaluated by measuring interpretation time per WSI. Differences in reading time between AI-assisted and unassisted conditions were analyzed using linear mixed-effects models, with review condition as a fixed effect and slide as a random intercept to account for repeated measurements. Statistical significance of fixed effects was assessed using Wald z-tests. Effect size for efficiency was quantified using Cohen’s d for paired samples, reflecting the within-subject crossover design.
4. **Preservation of ordinality**: We assessed the model’s ability to respect the continuity of histological grades (i.e., preserving the relationship Absent < Mild < Moderate < Marked). We performed slide-level embedding visualization, in which slide-level embeddings derived from the model were projected into a low-dimensional manifold (e.g., t-SNE). This allowed us to assess whether slides naturally clustered according to ordinal grades within each task.
5. **Visual interpretation**: We performed a qualitative assessment of the model’s interpretability by visualizing attention heatmaps at both the slide and patch levels. At the slide level, we aim to verify the MIL framework’s capacity to localize diagnostic regions by visualizing attention heatmaps, ensuring the model aligns with established topographical patterns of gastritis. Specifically, at the patch level, we will investigate the representational power of the Hibou-L encoder to determine if it captures the fine-grained morphological motifs required for identifying subtle microscopic entities. This includes validating whether the encoder can resolve *H. pylori* microorganisms—a task that typically necessitates 40x magnification— thereby confirming the model’s ability to ground global diagnostic decisions in high-fidelity, semantically meaningful evidence across different histological scales.
6. **Ablation test and baseline comparisons**: Finally, we conducted ablation studies to evaluate the sensitivity of the model to the hyperparameter *τ*, which controls the strength of the logit adjustment mechanism. To further evaluate the efficacy of our proposed framework, we compared SydneyMTL (utilizing logit adjustment loss) against two standard multi-task benchmarks: a baseline trained with Cross-entropy loss and a version utilizing Weighted Cross-entropy loss to account for class imbalance. All three models shared an identical attention-based Multiple Instance Learning architecture, ensuring that observed performance variances stemmed from the optimization objectives. We analyzed performance variations across different *τ* values to determine the optimal configuration for balancing ordinal constraints and classification accuracy.

### Implementation details

We processed the gigapixel WSIs into patches using a tessellation strategy. Specifically, a coarse tissue mask was generated on a downsampled image (5× magnification) using multi-level Otsu thresholding to distinguish tissue regions from the background. From the masked tissue regions, we extracted non-overlapping tiles measuring 224 x 224 pixels at 40x magnification. To exclude non-informative patches, we employed a JPEG compression-based filter; because JPEG efficiently encodes redundant low-frequency regions and homogeneous regions, such as backgrounds, and pen marks, it achieves higher compression for non-tissue areas, resulting in smaller file sizes.^29^ Consequently, patches with a JPEG file size below a predefined threshold (10 KB) were excluded. Each patch was then encoded into a 1024-dimensional feature vector using Hibou-L, a large-scale foundation model specifically pre-trained on diverse histopathology datasets.^30^

We implemented all deep learning models using the PyTorch framework. WSI preprocessing and tile management were executed using OpenSlide, OpenCV, and Pillow libraries. All training and evaluation procedures were conducted on a high-performance computing system equipped with 2x NVIDIA A100 GPUs (80 GB VRAM each) and an Intel Xeon Gold 6338N CPU (128 cores, 2.20 GHz) with 512 GB of RAM.

To optimize the network for clinical relevance, we projected the patch embeddings into a shared latent space via an adapter with a dimension of *d=*512. This configuration was selected through technical ablation studies to provide sufficient bandwidth for the joint detection of the ‘premalignant triad’ (Supplementary Table S1). The models were trained using the Adam optimizer with an initial learning rate 0.0001. To prioritize stable optimization and minimize gradient noise, we implemented a gradient accumulation strategy to achieve a larger effective batch size. Given that WSI bags are highly memory-intensive and exhibit significant variability in size, we utilized a physical batch size of one slide per iteration. By accumulating gradients over eight steps, we reached an effective batch size of 8, ensuring a robust training process while operating within hardware memory constraints. The training process was bounded to a maximum of 100 epochs. To prevent overfitting, we employed an early stopping mechanism: training was automatically halted if the validation loss did not show improvement for 10 consecutive epochs (patience=10).

## Ethics statement

All WSIs used in this study were obtained from Seegene Medical Foundation (SMF), Republic of Korea. These slides were originally collected during routine health checkups and external diagnostic consultations. The study was conducted in accordance with the principles of the Declaration of Helsinki and was approved by the Institutional Review Board of Seegene Medical Foundation (No. SMF-IRB-2024-015). All data were fully anonymized prior to analysis, and no identifiable personal information was accessed by the investigators. Given the retrospective nature of the study and the use of de-identified histological images, the requirement for informed consent was waived by the IRB.

## Resource availability

### Lead contact

Correspondence and requests for materials should be addressed to the lead contact, who will facilitate the fulfillment of such inquiries.

## Materials availability

This research did not produce any unique chemical reagents or biological materials.

## Data and code availability

The complete training and evaluation codebase is publicly available at GitHub (https://github.com/SMF-AI/SydneyMTL). While a public release is planned for the future, the anonymized Golden dataset (comprising 1024-dimensional feature embeddings extracted by Hibou-L, SydneyMTL predictions, and labels) is currently available upon reasonable request to the first or corresponding authors.

## Acknowledgements

None.

## Author contributions

Conceptualization, W.C.J, H.H.K, Y.S.K, and K.K; Data curation: W.C.J, H.H.K, Y.H, Y.S.K, and K.K; Methodology, W.C.J, and H.H.K.; Investigation, W.C.J, H.H.K, and G.H; Validation, W.C.J, H.H.K, Y.S.K, and K.K; Visualization, Y.H, W.C.J, H.H.K; writing—original draft, W.C.J and H.H.K.; writing—review & editing, W.C.J, H.H.K, and Y.S.K, and K.K

## Supplementary data

**Supplementary figure S1.**
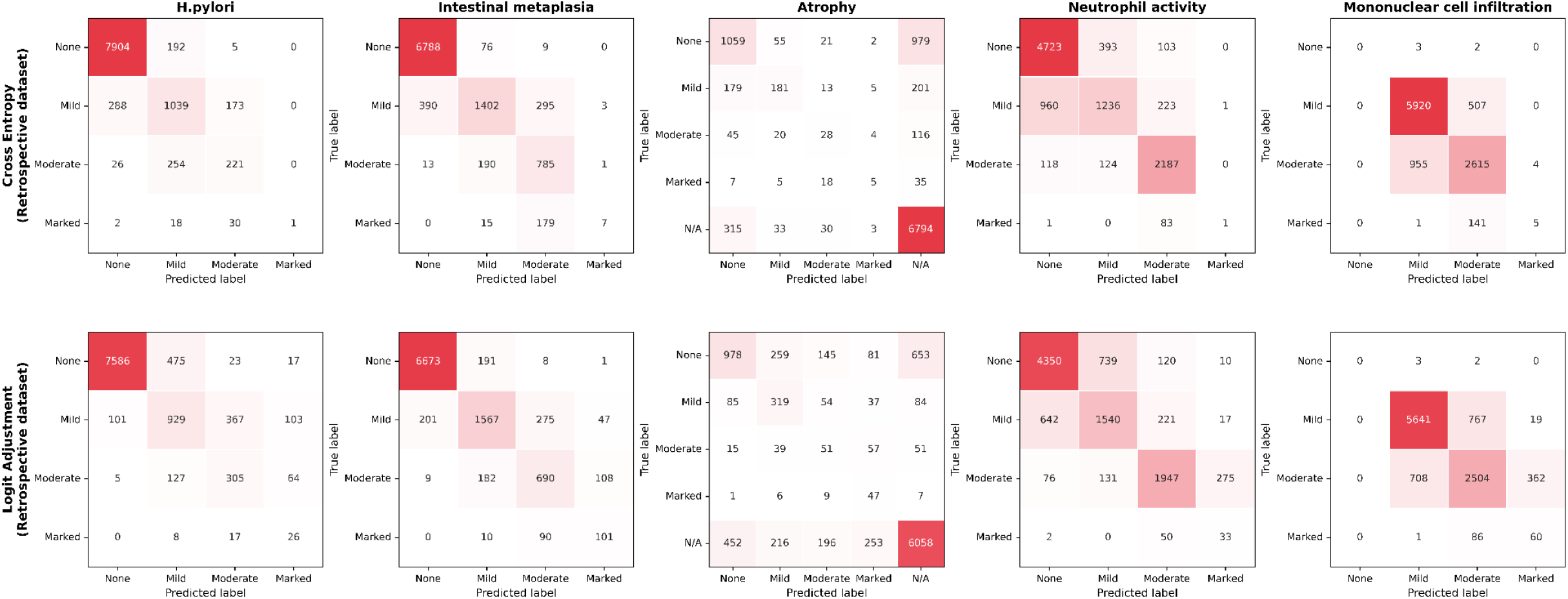
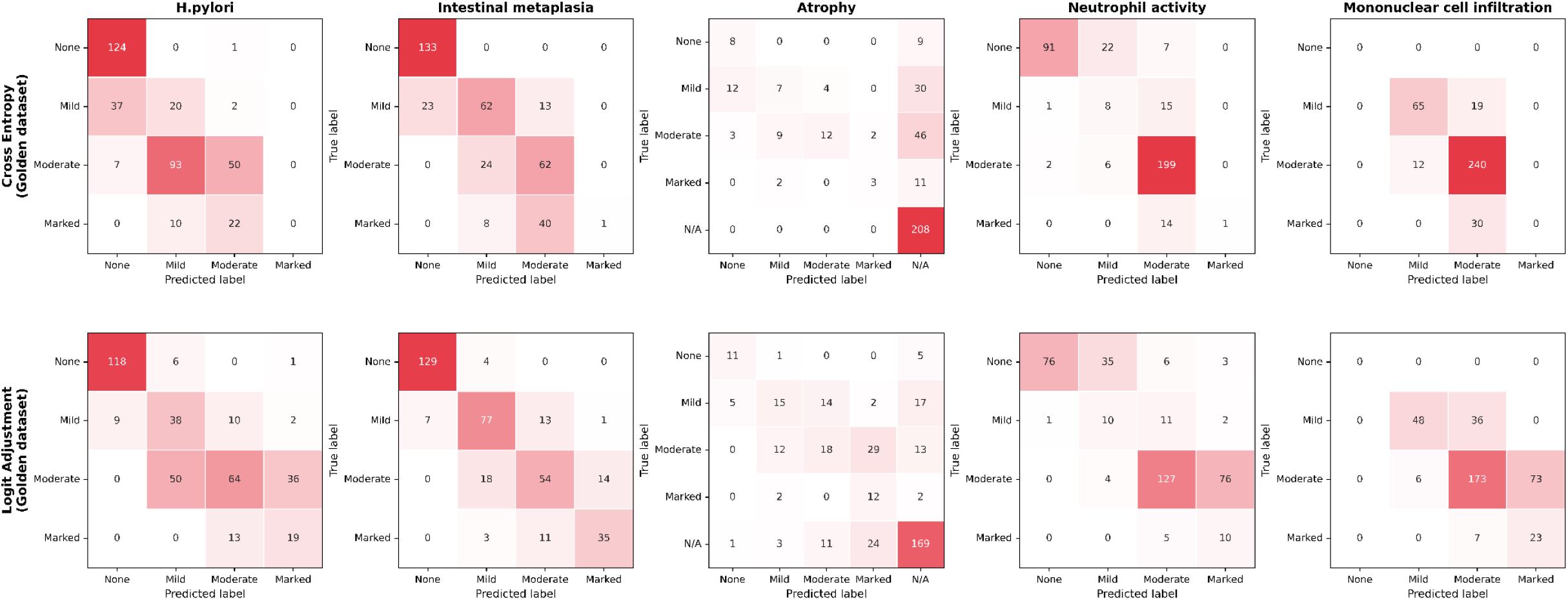
Confusion matrices of Cross-entropy and Logit Adjustment models on the Retrospective dataset and Golden dataset

**Supplementary figure S2.**
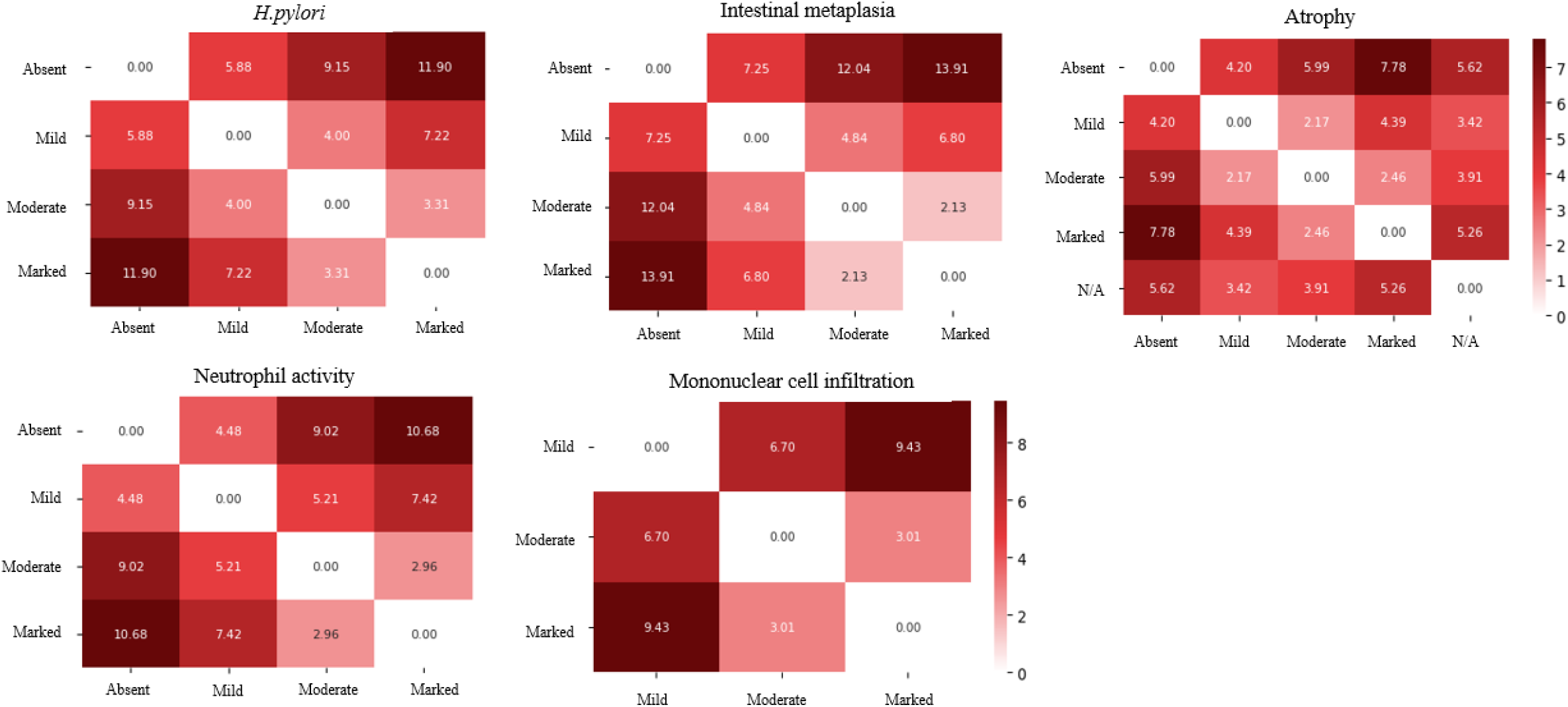
Quantitative validation of Ordinality via Inter-class Euclidean Centroid Distances. To substantiate the observed visual gradients, we calculated the distances between embedding centroids for each USS grade. Our analysis identifies a strict monotonic relationship, where distances increase proportionally with diagnostic divergence. Notably, for H. pylori, the distance from the “Absent” centroid scales linearly (5.88 for “Mild”, 9.15 for “Moderate”, and 11.90 for “Marked”), confirming that the model’s internal representation is governed by the underlying biological spectrum rather than discrete, unrelated categories.

**Supplementary figure S3.**
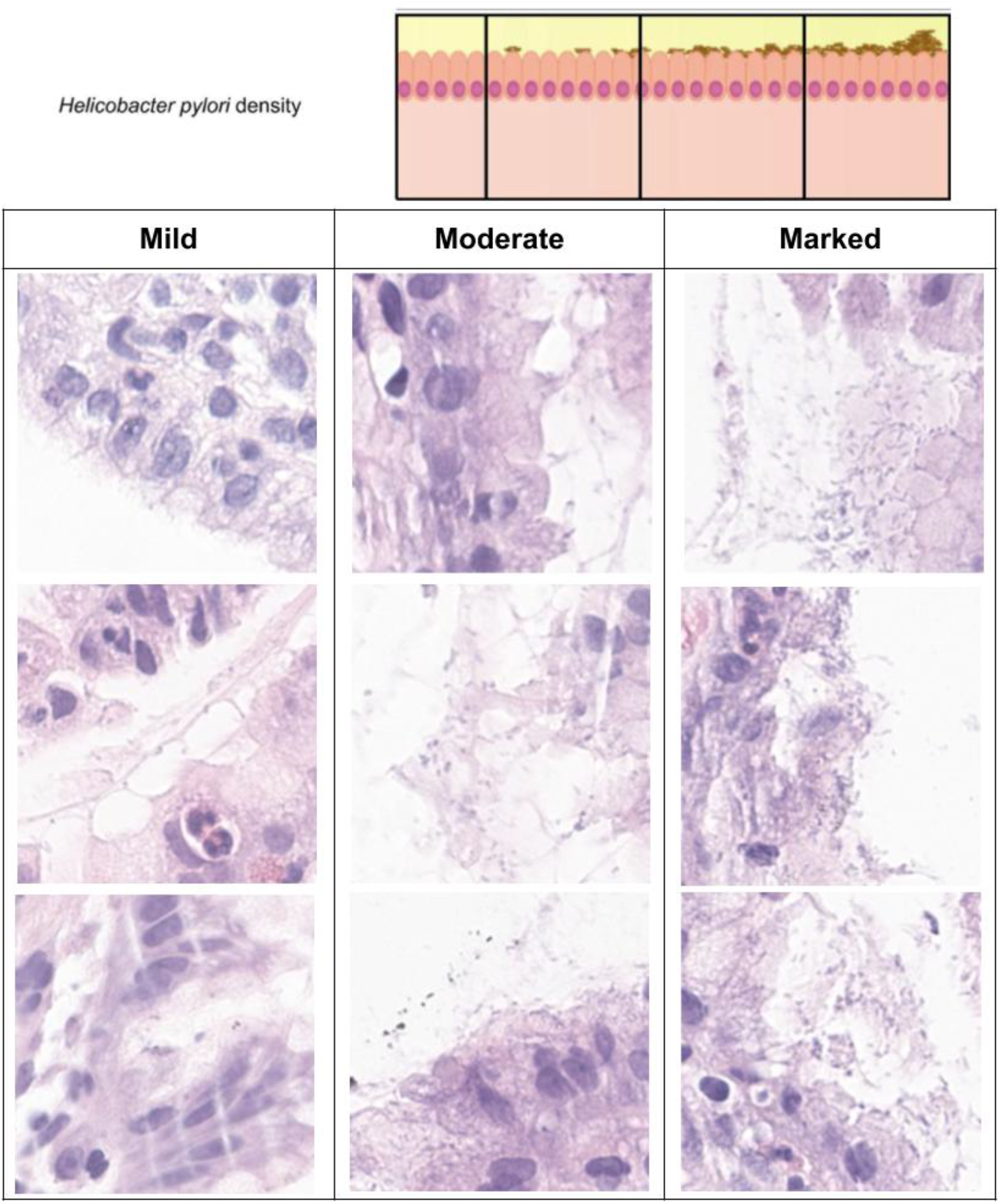
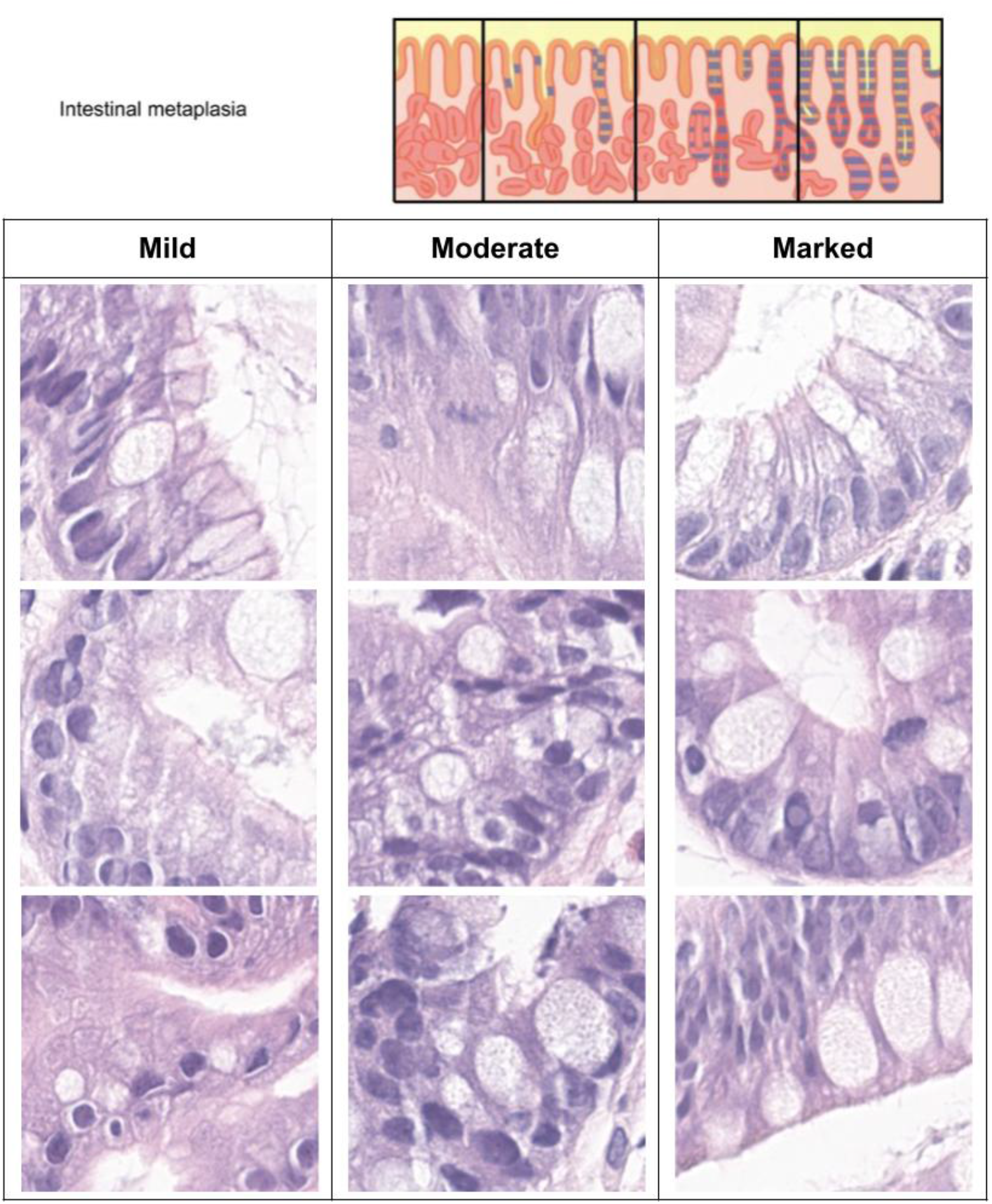
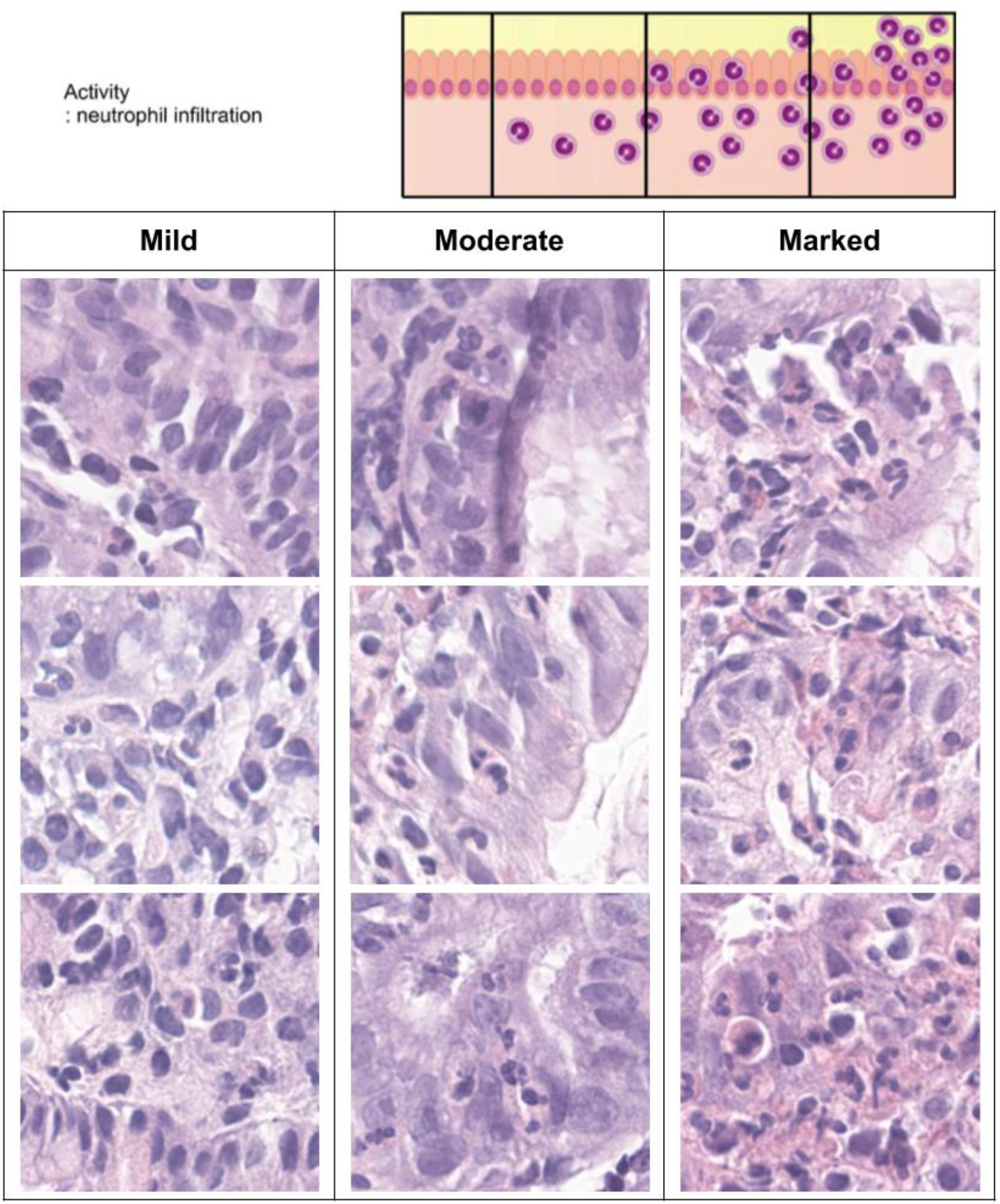
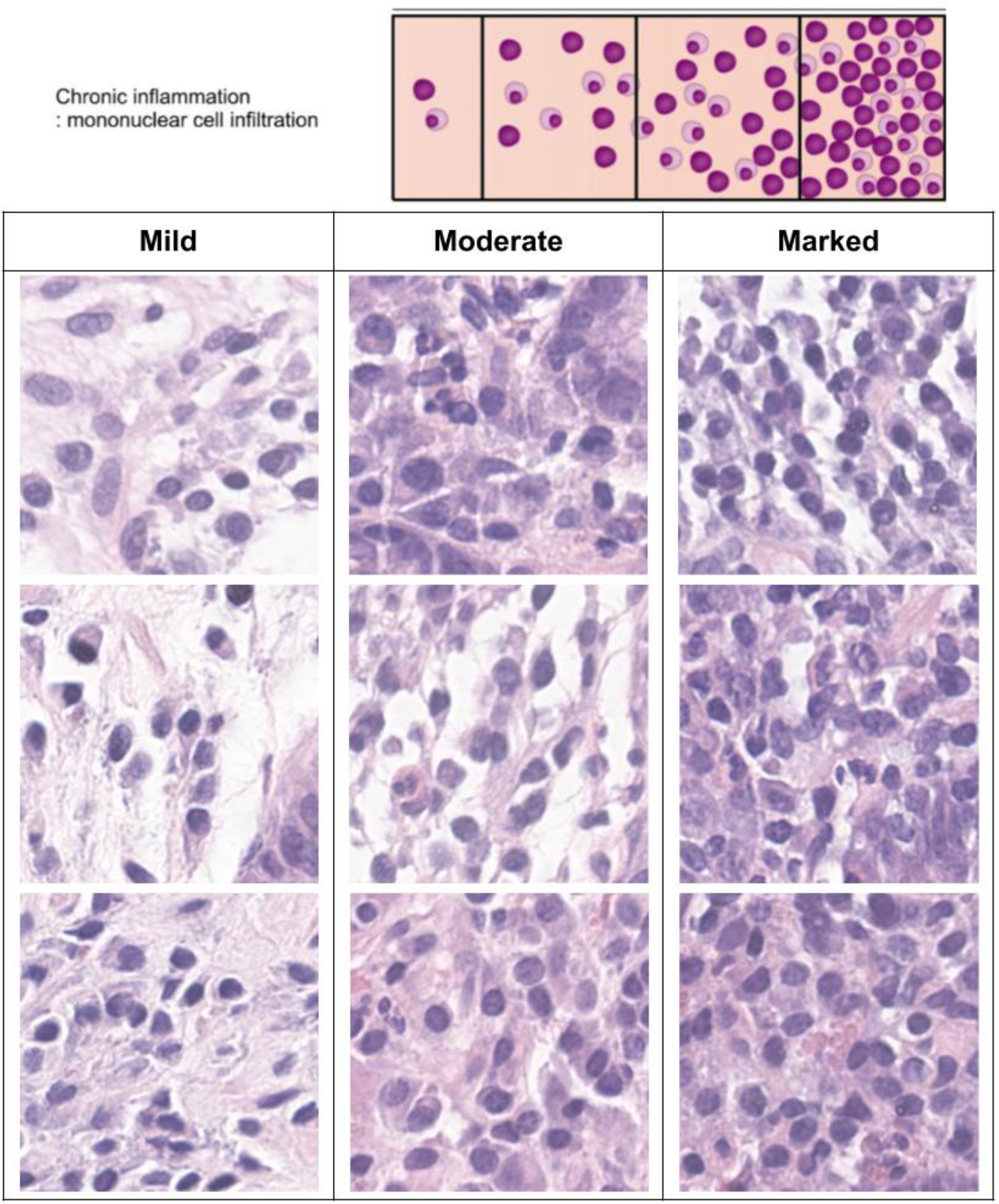
Visual analogues extracted from whole-slide images (WSIs) illustrating representative histologic appearances for each Sydney System attribute/grade

**Supplementary Table S1.**
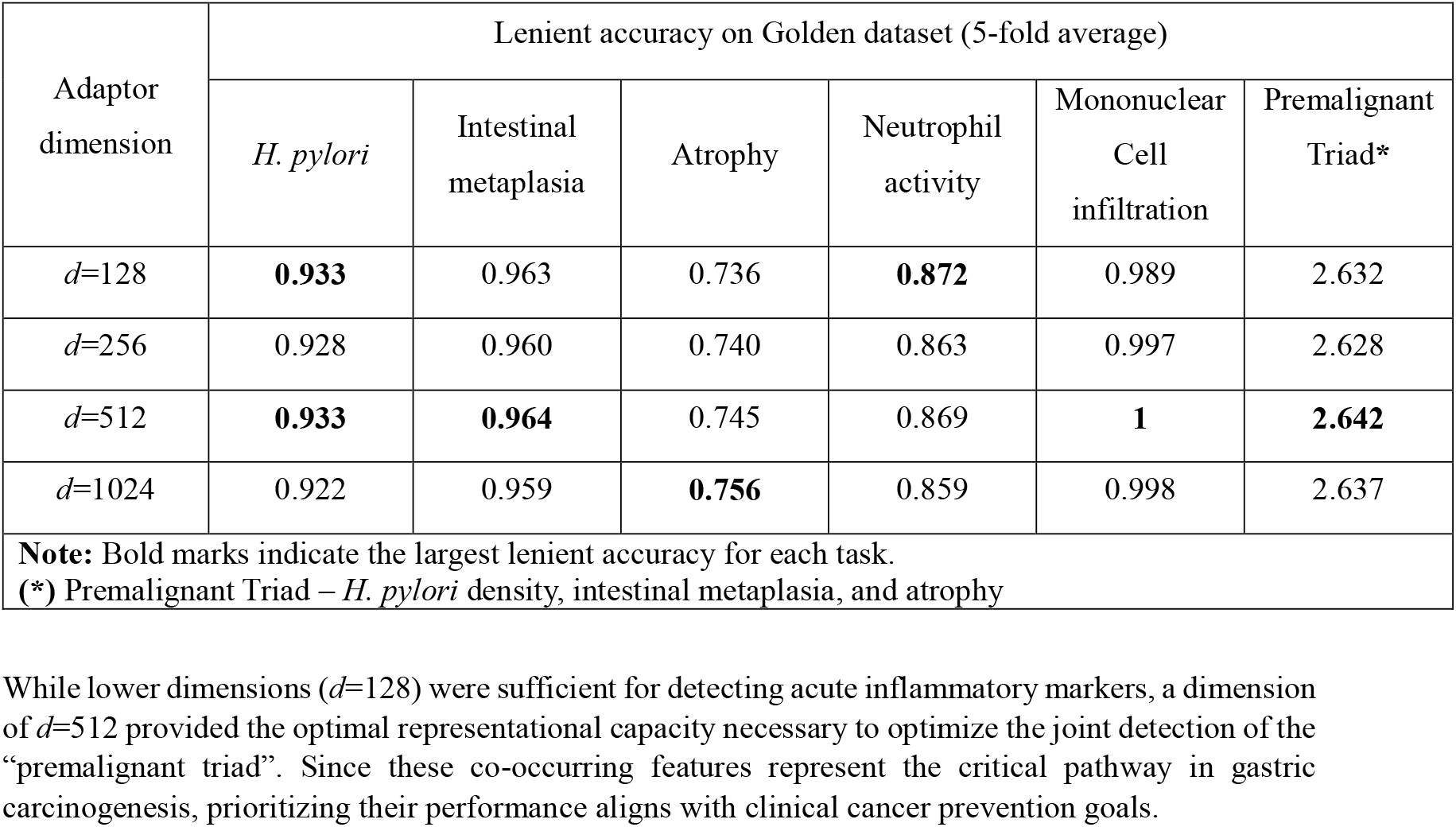
Ablation study of shared adapter dimension for clinical performance optimization.

